# Cumulus: A federated EHR-based learning system powered by FHIR and AI

**DOI:** 10.1101/2024.02.02.24301940

**Authors:** Andrew J. McMurry, Daniel I. Gottlieb, Timothy A. Miller, James R. Jones, Ashish Atreja, Jennifer Crago, Pankaja M. Desai, Brian E. Dixon, Matthew Garber, Vladimir Ignatov, Lyndsey A. Kirchner, Philip R. O. Payne, Anil J. Saldanha, Prabhu R. V. Shankar, Yauheni V. Solad, Elizabeth A. Sprouse, Michael Terry, Adam B. Wilcox, Kenneth D. Mandl

## Abstract

**Objective:** To address challenges in large-scale electronic health record (EHR) data exchange, we sought to develop, deploy, and test an open source, cloud-hosted app ‘listener’ that accesses standardized data across the SMART/HL7 Bulk FHIR Access application programming interface (API).

**Methods:** We advance a model for scalable, federated, data sharing and learning. Cumulus software is designed to address key technology and policy desiderata including local utility, control, and administrative simplicity as well as privacy preservation during robust data sharing, and AI for processing unstructured text.

**Results:** Cumulus relies on containerized, cloud-hosted software, installed within a healthcare organization’s security envelope. Cumulus accesses EHR data via the Bulk FHIR interface and streamlines automated processing and sharing. The modular design enables use of the latest AI and natural language processing tools and supports provider autonomy and administrative simplicity. In an initial test, Cumulus was deployed across five healthcare systems each partnered with public health. Cumulus output is patient counts which were aggregated into a table stratifying variables of interest to enable population health studies. All code is available open source. A policy stipulating that only aggregate data leave the institution greatly facilitated data sharing agreements.

**Discussion and Conclusion:** Cumulus addresses barriers to data sharing based on (1) federally required support for standard APIs (2), increasing use of cloud computing, and (3) advances in AI. There is potential for scalability to support learning across myriad network configurations and use cases.

## INTRODUCTION

The HITECH Act’s $48 billion federal investment led to the widespread adoption of electronic health records (EHRs), with over 95% uptake in both inpatient and outpatient settings.^1,2^ Though EHRs were not initially designed to support population level analytics or the exporting and sharing of data,^3^ they hold invaluable information that can be cornerstone assets for tasks requiring population data—tasks ranging from early warnings for emerging public health threats to training of artificial intelligence (AI) algorithms. The lack of standardization and technical complexities have traditionally made data extraction a challenge, restricting such capabilities to only the most technologically advanced healthcare systems with the time and resources to dedicate.

To facilitate EHR population data sharing at scale for public health purposes, we designed, developed, and tested Cumulus, a lightweight, open source and free, cloud hosted application that a provider organization can ‘plug in’ behind its firewall. The goals were to acquire, process and share population data on defined cohorts and enable ready participation in federated data sharing networks. To take advantage of free-text clinical notes data, Cumulus includes an AI natural language processing (NLP) pipeline to share information from unstructured data in a privacy preserving fashion with public health.

For nationwide scalability, Cumulus relies on data standardized in Fast Healthcare Interoperability Resources (FHIR) format and on a public application programming interface (API), SMART/HL7 Bulk FHIR Access.^4^ The API, which must be supported in all certified health information technology under the 21st Century Cures Act Rule,^5^ exposes as FHIR, the more than 100 data elements defined in the US Core for Data Interoperability (USCDI),^6^ which includes many categories of clinical notes.

Here we describe the Cumulus goals and architecture, and present early technical findings from its first deployment across five health systems, in partnership with state or local public health authorities.

## METHODS

Cumulus was developed under funding from the ONC Leading Edge Acceleration Projects program,^7^ instantiating 12 core technology and policy features to support health system-wide learning (Table 1).^8^ Though Cumulus is intended to multi-solve across many population health uses,^9^ the driving use case to inform its design was population health monitoring in collaboration between the care delivery system and public health agencies.

**Table 1.** Core technology and policy features instantiated in Cumulus software.

|  |
| --- |
| **Push button EHR data access.** Using the FHIR Bulk Data Access API, a healthcare institution can readily export data in a uniform, standardized format, eliminating complex extraction or mapping processes. |
| **Data processing pipeline.** Orchestration platform to automate end-to-end data extraction, de-identification, application of AI, aggregation, and transmission. |
| **Data de-identification.** Configurable data de-identification to minimize risk of unauthorized and unintended disclosures. |
| **Artificial intelligence / natural language processing of text.** Extraction of insights from clinical notes, converting them into FHIR data elements for privacy preserving sharing of insights from text. |
| **Provider site administrative simplification.** Integrated platform for end-to-end processing from EHR to public health and other use cases. |
| **Provider site autonomy.** Healthcare institutions remain in control of their EHR data and share when needed. |
| **Federated networking.** Uniform data export and sharing under the control of any healthcare institution (e.g., federally qualified health center, large academic health system) forms the basis of a de facto federated network. |
| **Advanced computable phenotyping.** Pre-configured AI/NLP algorithms for defining cohorts, endpoints, and outcomes from both structured and unstructured data. Enables dynamic cohort creation and data monitoring. |
| **Aggregate data sharing.** Preserves privacy by sharing only aggregate data counts (i.e., tallies) beyond the provider's firewall. |
| **Browser-based data access for third parties.** Enables authorized external parties to access aggregate data through dashboards and enclaves without the need for additional software. |
| **Turnkey deployment.** Simplified configuration of a cloud-hosted, containerized solution for rapid and secure installation behind a provider's firewall. |
| **Open source and free.** All Cumulus components are open source, liberally licensed, and available free of charge. |

### Design sprint

The project initiated with a five day, principled, user-centered design sprint following the Google Ventures method.^10,11^ The goal, in collaboration with public health, was to discover, plan and test features of a dashboard for public health practitioners. We defined goals, validated assumptions, and created wireframe prototypes, testing solutions with potential public health end users.

### Cloud Datastore

A common challenge in building and managing federated systems is deploying, maintaining and updating the infrastructure that runs at each node. In particular, tuning and scaling individual datastores as the amount of data increases accounts for a substantial component of the cost and complexity of these systems. To address this in Cumulus, we chose to leverage a managed cloud-based data lake environment as the primary data store, taking advantage of the scaling expertise of cloud providers to reduce the burden on individual institutions. We informally evaluated solutions from Amazon Web Services (AWS), Google Cloud, Microsoft Azure, and Databricks from a cost, functionality, and industry adoption perspective, ultimately deciding to use AWS Athena as a query engine over data stored in AWS S3 in DeltaTable format. This choice was heavily weighted by widespread existing use of AWS for clinical data in healthcare institutions at the time of selection. Longer term, we intend to develop software abstraction layers that can support the use of cloud data stores from other providers as well.

The landscape of components associated with the datastore such as de-identification and extract transform load (ETL) orchestration was similarly evaluated for the project. Only open source technologies were considered and key criteria included broad industry adoption and the ability to easily deploy them in the cloud and on-premises in Docker containers to enable institutions to configure their pipeline based on their organization’s preferences and policies. For example, some institutions preferred to de-identify data before transmitting it to a cloud storage bucket, while others preferred to run the de-identification process in their cloud provider. The initial Cumulus prototype used Apache Airflow as an ETL orchestration tool. Later versions moved to bespoke Python tooling for this purpose to increase development velocity.

### Modular AI/NLP

We performed a landscape analysis of clinical NLP tools for converting unstructured text in clinical notes to structured FHIR data serialized in JSON format. The NLP pipeline built on this work and was designed modularly to leverage rapidly evolving language models and always use the latest, validated models. Because we do not anticipate a workflow where providers commonly share the full text of clinical notes outside their institutions, our assumption was that NLP would occur behind the hospital firewall. Our initial work developing the SMART Text2FHIR pipeline demonstrates the feasibility of extracting privacy preserving standardized, structured FHIR data from notes before sharing.^12^

### Federated access

Recognizing that well-structured, queryable versions of clinical data are often unavailable or costly to obtain for care, research, or public health purposes, Cumulus was designed to function as a local environment in addition to a node in a federated network.

Additionally, this approach enables local users to leverage the NLP being applied to clinical notes to access data that would otherwise require substantial effort (e.g., chart reviews) to obtain. Using a local version of the Cumulus dashboard app, users can monitor key clinical metrics with minimal technical effort.

### Deployment and testing

In the context of a public health use case, the first Cumulus deployment was across five sites, one using a Cerner EHR, three using an Epic EHR, and one that implemented its own bulk FHIR Access API as a facade on top of a local data repository.

The Centers for Disease Control and Prevention contract supporting the work was administered through the CDC Foundation which interpreted federal requirements and classified the study as public health non-research that does not involve human participants and thus is exempt from human subjects research requirements. This determination was shared with the Boston Children’s Hospital Committee on Clinical Investigation, who concurred. The decision was communicated to all site principal investigators.

Example implementation metrics include time needed to configure Cumulus nodes and the scale of cohorts, encounters, and clinical notes able to be exported and processed during the pilot. Sites provided input on their experiences when implementing the open-source software, accessing bulk data through their FHIR APIs, discussions with EHR vendors, including any patches and updates to the API’s, and any local configurations they needed to make based on IT security or data sharing policies. We tested the distribution of a computable case definition across two sites. We also tested end-to-end dashboard access to aggregate data subscriptions by public health partners of two health care organization participants.

## RESULTS

### Architecture

The open source Cumulus system enables the dataflow shown in Figure 1. It is comprised of modular elements described here.

**Figure 1.**
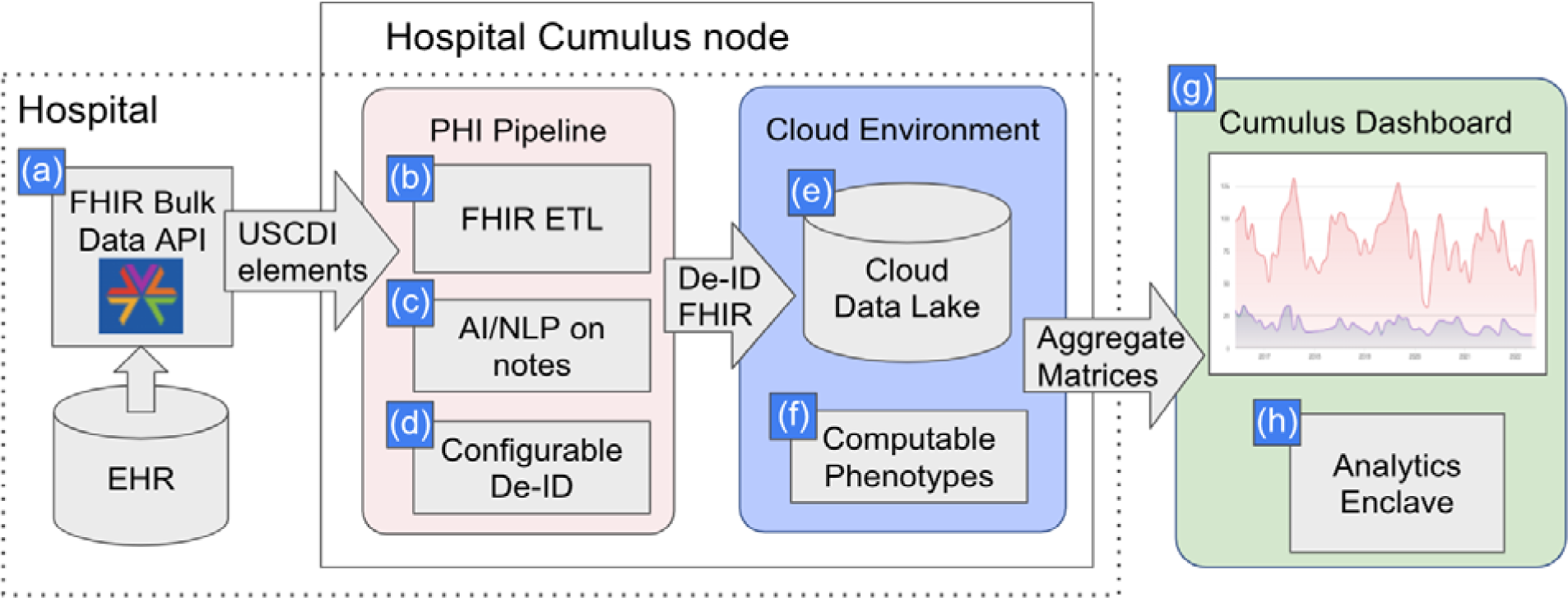
Cumulus Architecture Overview. We use Bulk FHIR **(a)** to retrieve from any electronic health record (EHR) all data elements contained in the US Core Data for Interoperability (USCDI). An extract, transform load (ETL) pipeline processes FHIR resources of potential interest to population health use cases and prepares them for analytic query **(b)**. Exported clinical notes are run through Natural Language Processing (NLP) **(c).** NLP-derived concepts and structured diagnoses, medications, and laboratory test results are then de-identified **(d)** before being uploaded to a locally controlled cloud-hosted data lake **(e)**. Computable phenotypes for the conditions of interest **(f)** are built from definitions in the open source Cumulus Library and used to screen the deidentified data for cases meeting the validated definitions. In the current configuration, aggregate matrices containing stratified variables of interest, such as new cases of COVID-19, are sent in a highly secure and privacy-preserving fashion to the graphically rich Cumulus dashboard **(g)**. Dashboard users can review and drill into the aggregate data and can perform their own analyses with the analytics enclave **(h)** using only a web browser.

### FHIR Bulk Data API

The ETL pipeline is a container run inside the health institution under local control. Within this container, a SMART/FHIR Bulk Data client authenticates with the local EHR and executes API queries to download USCDI data elements for cohorts of patients that are pre-defined in the EHR. Document references are parsed from this data and retrieved to obtain the raw text of clinical notes.

### NLP Pipeline

High throughput NLP methods, including large language models (LLM), are used to extract knowledge from clinical notes which is represented as JSON FHIR data models (resources). Each component of the NLP pipeline is instantiated as a container module (using Docker) with a REST interface for accessing separate functions. This modular design allows for multiple NLP pipelines to be run in parallel in the future. The default configuration runs the open-source Apache cTAKES, a library built for clinical text analysis that normalizes mentions of clinical entities to UMLS terminologies (SNOMED CT and RxNORM), and a BERT-based model for negation detection. The BERT-based model is trained on the same SHARP dataset used to train the cTAKES negation module,^13^ but updated with the latest pre-trained transformer-based methods.^14^ Because these modules are independent containers, each can be updated and re-deployed without changes to the client. NLP processing can occur under local control, behind a hospital firewall. No personally identifiable text from the notes is either retained or shared. Outputs of the NLP pipeline are saved to the cloud datastore as study variables for the aggregate matrix. To measure NLP accuracy, Cumulus library enables cohort selection to compare NLP results to human expert chart review. Cumulus enables chart review with an integration of LabelStudio, an open-source web-based tool for performing and managing manual data annotation. Clinical note storage, NLP processing, and chart review all occur within the control of the home institution. Currently, Cumulus supports notes presented in text or referenced as html documents. Additional note formats, such as PDF and XML, may be supported in the future.

### De-identification (DEID) pipeline

PHI is removed from structured FHIR data and FHIR data generated from the NLP pipeline prior to uploading to the cloud datastore. The open source, Microsoft FHIR anonymization tool^15^ is used to comprehensively remove unnecessary identifiers and only preserve de-identified fields of potential interest for population analysis. Cumulus provides templates for configuration of this tool, such that new health institutions can adopt standard practices that have been reviewed by multiple institutions. Additionally, a secure codebook method is used to obscure certain potentially identifying information such as patients’ unique FHIR identifiers from entering the local cloud data lake, which is under local control.

### Cloud Data Lake

The data lake and related cloud components can be configured on an AWS instance using CloudFormation templates. The de-identified data is loaded into an AWS S3 bucket in DeltaTable format and corresponding AWS Athena schemas are updated based on the FHIR data elements present in the data set.

### Cumulus Library

Recent improvements in storing and querying nested data with heterogeneous elements make modern cloud data stores ideal for processing FHIR data. However, native FHIR resources contain complex data structures that can make querying difficult, requiring the use of esoteric SQL features and resulting in very large queries. For this reason, Cumulus provides users with simplified, tabular representations of standardized FHIR data models covering visit data that includes patient demographics, vital signs, laboratory results, conditions and medications. Other data elements present in USCDI, such as allergies, immunizations, and implantable devices, can also be added to the library. Cumulus library also supports manual processes for importing existing standard value sets, such as the code lists publicly available in the National Library of Medicine Value Set Authority Center (VSAC).

Study criteria contain three parts: case definition, study variables, and the study period. Each study selects one or more patient cohorts. By default, only patients matching the case definition in the study period are selected. Optionally, propensity score matching is used to select cohorts for comparison. Cumulus library uses the study criteria to calculate an aggregate matrix containing counts of all study variables for each defined patient cohort.

### Computable phenotypes

A central feature is the ability to collaboratively define computable phenotypes or computable case definitions to identify diverse, representative patient groups using either USCDI coded data alone or incorporating structured data, optionally enhanced by NLP of text. Several clinical domains were explored. Symptoms of COVID-19^16^ were NLP extracted and output as FHIR resources. Cumulus provides a powerful library that combines the simplicity of SQL with the quality of FHIR resources extracted from the longitudinal patient history. The broad range of needs for computable phenotypes resulted in Cumulus support for AI/NLP, standard VSAC value sets, and custom user defined criteria. For example, the Cumulus hypertension definition is based on the CMS electronic quality measure that provided inclusion and exclusion criteria for hypertension treatment.^17^ Computable phenotypes were derived from FHIR observation vital signs at critical values of 140/90 mm/hg. Custom value sets were user defined for self-harm in the domains of mental health study^18^ and opioid overdose.

### Aggregate matrix

Counts of every study variable combination are calculated first by each participating healthcare site and then aggregated to produce a sum total of counts across the Cumulus network for the Cumulus Dashboard and Analytics Enclave. The aggregate matrix can be refreshed on demand or as a scheduled task depending on the clinical study needs. The subscription metadata includes which healthcare sites provided data, the study period of the data collection, descriptions of study variables, and when the aggregate matrix was compiled. Formally, the aggregate matrix is a power set^19^ containing counts of all combinations of study variables, including the null set representing the total size of the selected cohort (Equation 1). The Cumulus library produces the aggregate matrix by generating a SQL select count query with the cube^20^ function. The user specifies the study variables to count, typically the number of patients or encounters. The cardinality of the power set is 2^*n,* where *n* is the number of discrete elements among the study variables. In practice, the aggregate matrix is much more sparse than a pure power set: Cumulus removes set sizes with fewer than 10 patients. Examples of discrete elements include disease status (Boolean), patient age at encounter (integer), encounter month (date), and antihypertensive medications.^21,22^.

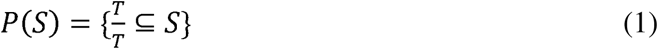

**Equation 1**. Cumulus aggregate matrix is a power set. Let *S* be a set. *T* represents a subset of *S.* P(*S*) denotes the power set of *S* as all subsets of *S*, including the empty set and *S* itself. The discrete elements of each subset *T* include one or more study variables.

### Cumulus Dashboard

The design sprint prototype instantiated key features including facilitated iterative data exploration and refinement of definitions, user review of freshness and provenance of data in the dashboard for context, power user analyses on the data *without downloading or managing additional software*, and a workflow for requesting targeted exports of line-level data for vetted use cases. Development of the dashboard is ongoing with input from users. The Cumulus Dashboard provides public health users with the ability to graph, stratify, and compare patient populations using any combination of requested study variables from the aggregate matrix. Users select the graph type that best represents the relationship to visualize: line graph, bar chart, pie chart, or area chart. Users apply filters to include or exclude patient populations, for example filtering by age at encounter, encounter week, diagnosis, test result, or any other study variable. The user selects which study variables to graph and whether to present counts or percentages of the population. Figure 2 is an example of COVID-19 symptoms graphed by encounter month. It shows the prevalence of patient symptoms at emergency department encounters during the COVID-19 pandemic. In this example, the dashboard is used to compare two methods for measuring symptoms, NLP computable phenotypes and ICD-10 diagnosis codes.^16^

**Figure 2.**
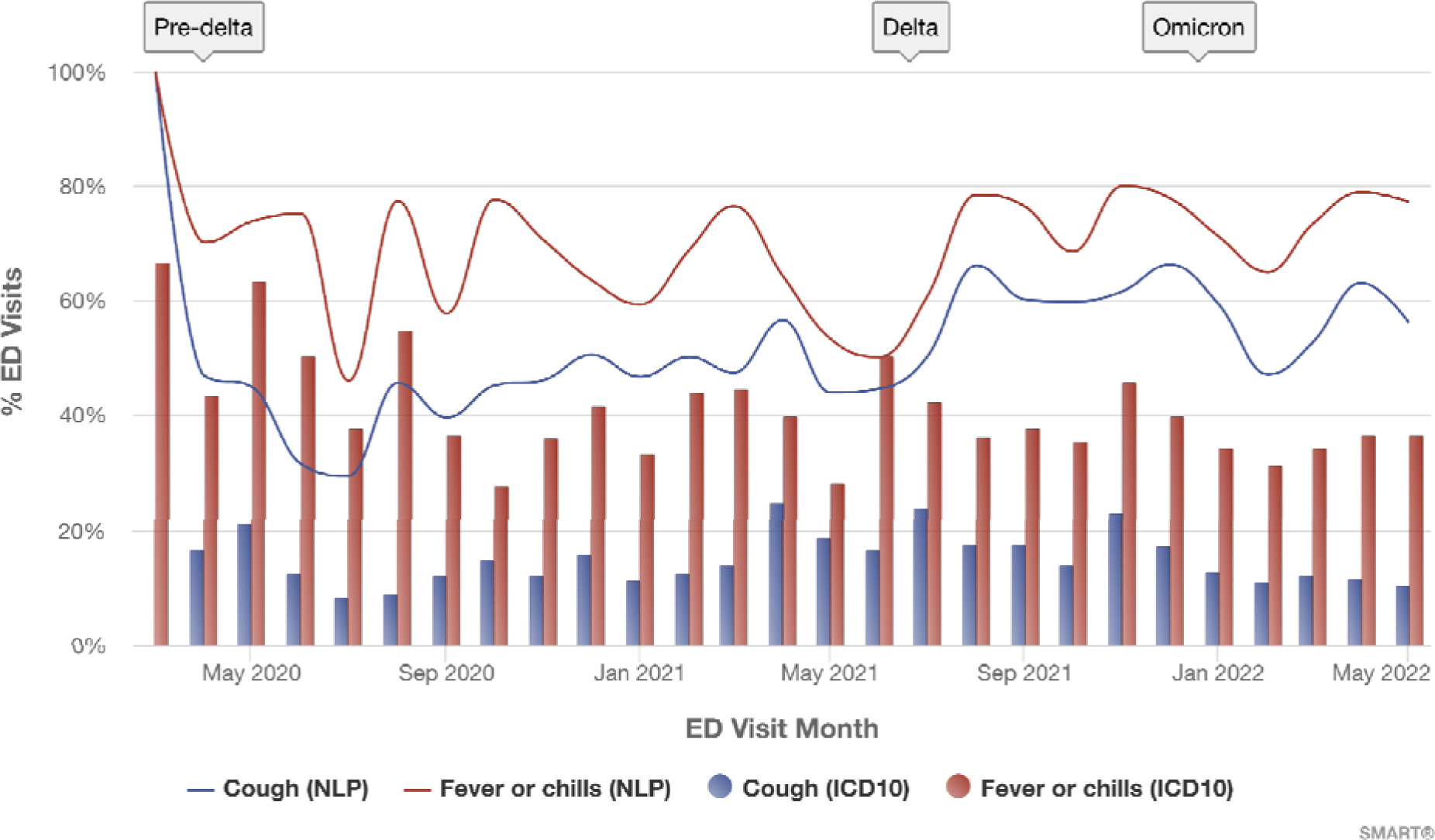
Cough and fever symptom trends for patients with COVID-19 visiting the emergency department (ED) between March 2020 and May 2022, as identified using two methods. Lines represent the percent of COVID-19 patients with each symptom using NLP-driven computable case definitions. Bars represent the percent identified using ICD-10 codes. Red denotes fever or chills, blue denotes cough. Symptoms were detected in a greater percentage of ED visits for patients with COVID-19 using NLP versus ICD-10.

### Analytics enclave

The enclave, for power users to programmatically analyze Cumulus aggregate data, is a Python notebook accessed via a web browser. The Python notebook is preloaded regularly with Cumulus aggregate data and commonly-used data science software for graphing and numerical analysis (Matplotlib, Pandas, SciPy, scikit-learn, others). Cumulus aggregate data include all combinations of study variables as a power set and crosstab tables. These counts are directly applicable for population health measures including disease prevalence, odds ratio, relative risk, conditional probability, chi-square tests for significance, Bayesian classifiers, and decision tree classifiers. The analytics environment also enables propensity score matching.

### Federation

The Cumulus network implements the federalist principles of local control for healthcare sites,^23^ privacy protection for patients, and sharing of aggregated counts for authorized users. Cumulus federation is a push model and not a query model—no central authority has access to directly query the line-level data of any participating institution. The aggregate matrix includes every pre-computed combination of study variables, allowing for near-instantaneous responses to user actions. A Cumulus network deployed with five sites—four hospitals and one health information exchange (HIE)—is shown in Figure 3. Each site remains in control of the patient data they are legally responsible for as a HIPAA covered entity. The ETL pipeline is a container within the site intranet behind the firewall that extracts data from EHR, runs NLP and de-identification pipelines, and loads the prepared FHIR data into the private cloud environment. Cumulus library is run within the private cloud and outputs an aggregate matrix of counts for each study. Each site uploads the aggregate data to a coordinating site. The aggregate matrix is then merged across all five sites resulting in a sum total of counts across the network. The aggregated dataset only contains counts. Credentialed users are then able to graph and analyze the aggregate data using the Cumulus Dashboard and Analytics Enclave.

**Figure 3.**
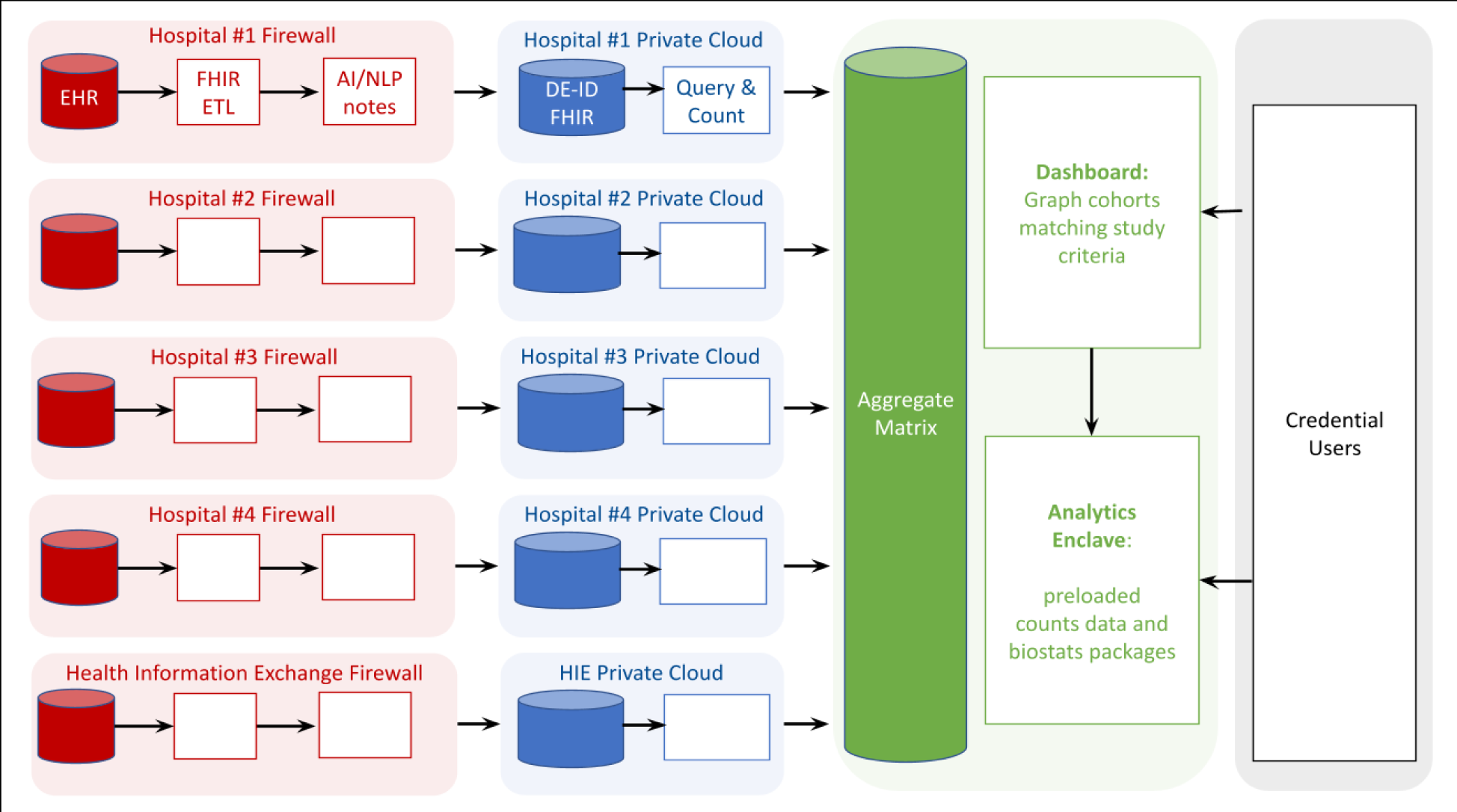
Cumulus configured within a federated network of five healthcare organizations (four academic medical centers and one health information exchange). The left (red) column denotes data behind each hospital firewall, where FHIR coded data are extracted and natural language processing performed to extract computable case criteria from patient notes. The next (blue) column denotes de-identified data in private clouds with elements of personally identifiable information removed. The next (green) column denotes aggregated patient counts shared with authorized public health users (final, right-side column). Empty boxes denote the same configurations as Hospital #1, except that in the left (red) column, only the four hospitals used FHIR ETL to connect to their local EHR.

### Deployment

The first Cumulus testbed was deployed at five dyads of healthcare organizations with public health partners: Boston Children’s Hospital (BCH) and Massachusetts Department of Public Health, Regenstrief Institute and Marion County Public Health Department, Rush University Medical Center and Chicago Department of Public Health, Washington University in St. Louis and City of St. Louis Department of Health, and UC Davis Health and Yolo County Health and Human Services and Sacramento County Public Health.

There were some local variations in Cumulus installations. Regenstrief Institute developed a bulk FHIR API following the Bulk Data Implementation Guide to surface USCDI elements from an existing data warehouse containing HL7 V2 messages stored by the Indiana Health Information Exchange. University of California Davis Health installed the Cumulus ETL and NLP pipelines on premises to connect through a locally required API manager service rather than in the cloud. Washington University in St. Louis ran the ETL and NLP pipelines in a locally approved Microsoft Azure environment. All sites configured and ran the Cumulus Library queries in AWS and sent aggregate results to BCH for the dashboard. Features of the implementation are captured in Table 2.

**Table 2.** Implementation statistics for five sites.

|  | **University of California, Davis Health** | **Regenstrief Institute** | **Boston Children's Hospital** | **Rush University Health** | **Washington University of St. Louis** |
| --- | --- | --- | --- | --- | --- |
| Unique FHIR patient records loaded | 222 (+12k soon) | 334,573 | 179,176 | 2,001 | 1,268 |
| FHIR Encounters loaded | 6K (+4.45M soon) | 9.2M | 3M | 385K | 0 (250K soon) |
| Clinical notes processed | 23K (+6.25M soon, metadata only) | 23.2M | 1.8M | 157K (metadata only) | 0 (328K soon) |
| Symptom mentions extracted via computable case definition |  | 609,833 | 85,678 |  |  |
| FHIR API vendor | Epic | Custom HIE API | Cerner | Epic | Epic |
| PHI processing environment | on-prem | on-prem | AWS | AWS | Azure |
| Configuration time after approvals | 19 calendar days | 53 calendar days | 14 calendar days | 26 calendar days | 39 calendar days |
| API supported date filtering? | N | Y | Y | N | N |
| Health Departments with live dashboard in test bed | Yolo County Health and Human Services & Sacramento County Public Health | Marion County Public Health Department |  |  |  |

An early measurement made in the testbed was performance of the first nascent implementations of the Bulk FHIR API across multiple dimensions.^24^ The first wave of APIs varied across vendors, products, and configurations, from the order of 2,000 to 11,000 FHIR data models (FHIR Resources) per minute. Sites with access to APIs that supported optional date filtering parameters were able to make more targeted requests and could export relevant data for larger cohorts of interest. It was observed that APIs with low throughput compared to their patient volume had to restrict inclusion criteria to complete exports of USCDI elements within their EHR. Additionally, APIs that did not support date filtering were unable to efficiently refresh cohort data with FHIR resources created after a previous export; the entire patient history had to be exported for each request.

## DISCUSSION

The Cumulus cloud hosted ‘listener’ is a viable technology to instrument provider sites to access data from EHRs for public health purposes or internal uses. It functions as an ‘app’ running against the universally available SMART/HL7 Bulk FHIR Access API—now required by the 21st Century Cures Act—that facilitates point to point data exchange of both structured and free text data.

We demonstrated the capacity for Cumulus to serve as a node in a federated network. In this architecture, Cumulus shares insights while only exchanging deidentified aggregate data outside the healthcare institution. In future work, line level extracts could be provided as a second step in public health or other workflows, with consideration to all required data access policy and legal implications. We introduced this approach for a national information infrastructure demonstration,^25^ used it extensively in the SHRINE network,^26,27^ with disease registries,^28^ the current NCATS-funded Genomic Information Commons (Mandl PI),^29^ and it has become the model for our collaborators establishing PCORNet capabilities.^30^

By ensuring that institutions catering to the full spectrum of patient demographics—from major medical centers to federally qualified health centers—can participate in public health surveillance, we aim to achieve more equitable, inclusive, and representative data. This approach is crucial for public health response, developing treatments, and implementing healthcare strategies that are effective across populations, irrespective of their ethnic, socio-economic, or geographical backgrounds.

We identified key technical challenges. The AI/NLP/LLM field is progressing at an unprecedented pace, posing a challenge to keep the Cumulus project aligned with the latest advancements. We plan to address this by implementing robust, reproducible, and reusable benchmarking and validation routines. Modular interfaces to the AI model are used to minimize hard-coupling. These routines are specifically designed to be adaptable and effective for emerging models, ensuring that Cumulus remains at the forefront of technological advancements. Because the output and quality of LLM models can vary significantly over time, impacting the consistency and reliability of the phenotyping, we will investigate open source tooling in the ecosystem to monitor LLM quality and validity.

Currently, there is not a widely adopted, standardized set of metrics to assess the data quality of FHIR elements extracted from an EHR making it unclear what level of quality a healthcare institution or public health organization can expect when using these datasets. Future work will address this by defining a broad set of FHIR data quality metrics focused on the USCDI dataset in collaboration with a range of interested parties, developing open source tooling to execute these metrics at care delivery sites using the Cumulus infrastructure, and sharing benchmarks of the results from pilot sites.^7^ This work will support the access of standardized data within EHRs.

The only data to leave the health care site are aggregated counts (tallies) of the number of patients matching case criteria. Further, patient counts are only shared with credentialed end users, for example with a known clinical research study or public health use case. If security were ever compromised, patient counts do not constitute a HIPAA disclosure and do not require patient contact. This greatly simplifies administrative and IT security reviews, lowering the barrier to participation.

## CONCLUSION

Cumulus tackles obstacles to data sharing through mandated support for standard APIs, the growing adoption of cloud computing, and advancements in artificial intelligence. This approach offers the scalability needed to facilitate learning across various self-organized federated network configurations and use cases.

## DATA AND CODE AVAILABILITY

Data and code are available at https://docs.smarthealthit.org/cumulus/

ETL Pipeline is available at https://github.com/smart-on-fhir/cumulus-etl

Library is available at https://github.com/smart-on-fhir/cumulus-library

Computable case definition libraries for COVID-19 symptoms, hypertension, suicidality, and opioid overdose and use disorder are available at:

https://github.com/smart-on-fhir/cumulus-library-covid https://github.com/smart-on-fhir/cumulus-library-hypertension/ https://github.com/smart-on-fhir/cumulus-library-suicidality-los https://github.com/smart-on-fhir/cumulus-library-opioid

Aggregator is available at https://github.com/smart-on-fhir/cumulus-aggregator

## AUTHOR CONTRIBUTIONS

KDM obtained funding. AM, DG, TM, JRJ, ES, KDM conceptualized the study and wrote the first draft. AM, JRJ, AA, JC, PMD, BD, MG, VI, L**A**K, PR**O**P, AS, P**RV**S, **YVS,** ES, MT, ABW were involved in data curation and project administration. MG, VI, MT developed the software. DG, JRJ, BD, LK, PS, ES, PRP, KDM conducted the formal analysis. All authors were involved in review and editing.

## ACKNOWLEDGEMENTS

This work was supported by: The Office of the National Coordinator of Health Information Technology contract numbers 90AX0031/01-00, 90AX0022/01-00, and 90AX0040/01-00; Centers for Disease Control and Prevention of the United States Department of Health and Human Services (HHS) as part of a financial assistance award, Strengthened Community Partnerships for More Holistic Approaches to Interoperability totaling $1,985,178. The contents are those of the author(s) and do not necessarily represent the official views of, nor an endorsement, by the CDC Foundation, CDC/HHS, or the U.S. Government; The National Center for Advancing Translational Sciences/National Institutes of Health Cooperative Agreements U01TR002623 and U01TR002997; National Association of Chronic Disease Directors/Centers for Disease Control and Prevention Grant No. NU38OT000286; Centers for Disease Control and Prevention Grant No. U18DP006500; Centers for Disease Control and Prevention Cooperative Agreement No. NU58IP000004, 1U01TR002997-01A1.

## COMPETING INTERESTS

Boston Children’s Hospital receives philanthropic contributions on behalf of the laboratory of K.D.M. from the SMART Advisory Committee with members including Microsoft, Cambia, Humana, and HCA Healthcare.

